# Investigation of *RFC1* tandem nucleotide repeat locus in diverse neurodegenerative outcomes in an Indian cohort

**DOI:** 10.1101/2023.06.05.23290839

**Authors:** Nishu Tyagi, Bharathram Uppili, Pooja Sharma, Shaista Parveen, Sheeba Saifi, Abhinav Jain, Akhilesh Sonakar, Istaq Ahmed, Shweta Sahni, Uzma Shamim, Avni Anand, Varun Suroliya, Vivekanand Asokachandran, Achal Srivastava, Sridhar Sivasubbu, Vinod Scaria, Mohammed Faruq

**Affiliations:** Genomics and Molecular Medicine Division, CSIR - Institute of Genomics and Integrative Biology, New Delhi, 110007, India; Academy of Scientific and Innovative Research (AcSIR), Ghaziabad, 201002, India; Department of Neurology, Neuroscience Centre, All India Institute of Medical Sciences (AIIMS), 110608, New Delhi, India

**Keywords:** Ataxia, IndiGen, *RFC1*, Tandem nucleotide repeat expansion, Haplotype, CANVAS, SCA12, *Alu*Sx3

## Abstract

**Background and Objectives:** An intronic bi-allelic pentanucleotide repeat expansion mutation, (AAGGG)_400-2000_ at AAAAG repeat locus in *RFC1* gene is known as underlying genetic cause in cases with cerebellar ataxia, neuropathy and vestibular areflexia syndrome (CANVAS) and late onset sporadic ataxia. Biallelic positive cases carry a common recessive risk haplotype, ‘AAGA’ spanning *RFC1* gene. In this study our aim is to find prevalence of bi-allelic (AAGGG)_exp_ in Indian ataxia and other neurological disorders and investigate the complexity of *RFC1* repeat locus and its potential association with neurodegenerative diseases in Indian population-based cohorts.

**Methods:** We carried out repeat number and repeat type estimation using flanking PCR and repeat primed PCR (AAAAG/AAAGG/AAGGG) in three Indian disease cohorts and healthy controls. Haplotype assessment of suspected cases was done by genotyping and confirmed by sanger sequencing. Blood samples and consent of all the cases and detailed clinical details of positive cases were collected in collaboration with AIIMS. Further, comprehension of *RFC1* repeat locus and risk haplotype analysis in Indian background was performed on the NGS data of Indian healthy controls by ExpansionHunter, ExpansionHunter de novo and PHASE analysis respectively.

**Results:** Genetic screening of *RFC1-*TNR locus in 1998 uncharacterised cases (SCA12: 87; Uncharacterised ataxia: 1818, CMT: 93) and 564 heterogenous controls showed that the frequency of subjects with bi-allelic (AAGGG)_exp_ are 1.15%, <0.05%, 2.15% and 0% respectively. Two *RFC1* positive sporadic late onset ataxia cases, one bi-allelic (AAGGG)_exp_ and another, (AAAGG)_exp_/(AAGGG)_exp_ had recessive risk haplotype and CANVAS symptoms. Long normal alleles, 15-27, are significantly rare in ataxia cohort. In IndiGen control population (IndiGen; N=1029), long normal repeat range, 15-27, is significantly associated with A_3_G_3_ and some rare repeat motifs, AGAGG, AACGG, AAGAG, and AAGGC. Risk-associated ‘AAGA’ haplotype of the original pathogenic expansion of A_2_G_3_ was found associated with the A_3_G_3_ representing alleles in background population.

**Discussion:** Apart from bi-allelic (AAGGG)_exp_, we report cases with a new pathogenic expansion of (AAAGG)_exp_/(AAGGG)_exp_ in *RFC1* and recessive risk haplotype, ‘AAGA’. We found different repeat motifs at *RFC1* TNR locus, like, AAAAG, AAAGG, AAAGGG, AAAAGG, AAGAG, AACGG, AAGGC, AGAGG, AAGGG, in Indian background population except ACAGG and (AAAGG)_n_/(AAGGG)_n_. Our findings will help in further understanding the role of long normal repeat size and different repeat motifs specifically, AAAGG, AAAGGG and other rare repeat motifs, at *RFC1* locus.

Graphical Abstract:
Genetic heterogenicity at *RFC1*-TNR locus.

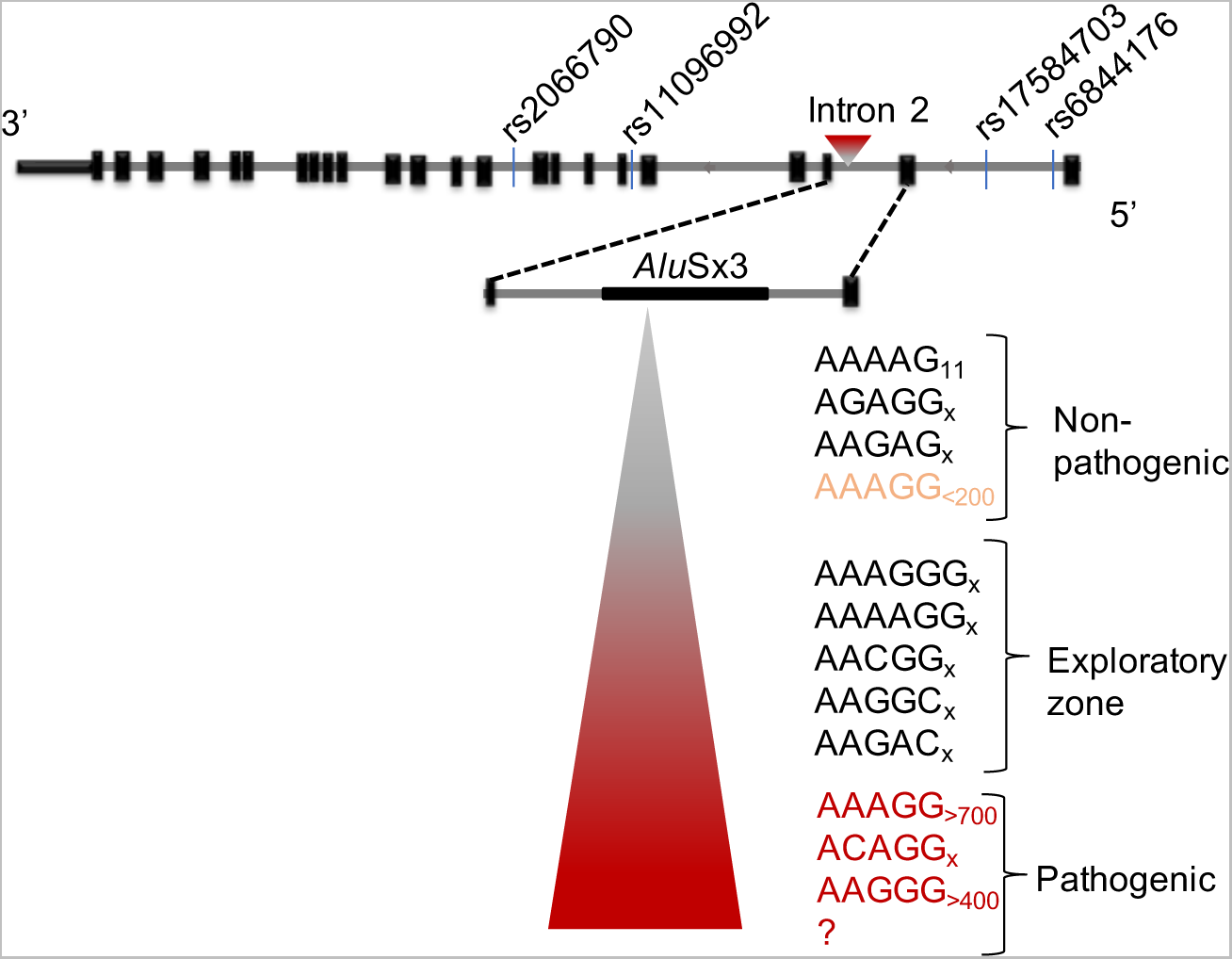

## Introduction

Short tandem repeat expansions are known to cause some of the spinocerebellar ataxias (SCAs), dentatorubral-pallidoluysian atrophy (DRPLA), Friedreich’s ataxia (FRDA), and the cerebellar ataxia, neuropathy, and vestibular areflexia (CANVAS) syndrome. As of today, more than 50 hereditary neurological, neuromuscular, and other neurodevelopmental disorders have been linked to TNR expansions in promoters, non-coding regions, and coding regions. ‘CAG’ repeat expansion, as in SCA1, SCA2, SCA3, SCA6, SCA7, SCA12, and SCA17, is the most common STR expansion in patients with inherited cerebellar ataxia. SCA1, SCA2, SCA3, and SCA12 are the most prevalent forms of autosomal dominant ataxia Compared to SCA6, SCA7, SCA8, SCA10, and SCA17 in India^1^. FRDA is the most common autosomal recessive ataxia, followed by Ataxia Telangiectasia, Spastic Ataxia of Charlevoix-Saguenay, and Ataxia oculomotor apraxia type 2^2–4^. Several investigations demonstrate that SCA1, 2, 3, 12, and FRDA share a common founder in the Indian population^5^. Among comparison to other parts of the world, SCA12 is more prevalent in the Indian population^6,7^. 60-70% of ataxia cases worldwide still lack genetic characterization and same has been seen in the Indian ataxia cohort according to Sharma et al., 2022^4^. In 2019, Cortese et al found a new biallelic penta-nucleotide (AAGGG)_400-2000_ recessive repeat expansion in poly-A tail of *Alu*Sx3 element present in intron 2 of *RFC1*^8^. Although this repeat locus in the reference genome contains (AAAAG)_11_ pure repeats, it is also prone to have other repeat motifs, AAAGGG, AACGG, AAGGC, AACGG, AAAGG, AAGAG, AGAGG, and ACAGG other than AAAAG and AAGGG^9–12^. Genetic testing has identified the pathogenic variant (AAAGG)_10-25_(AAGGG)_exp_ in New Zealand Maori and Cook Island Maori CANVAS patients^13^. A clinical CANVAS case with STR alleles having the unique repeat motifs (AAGGG)_1010_ and (AAAGG)_960_ has been found^14^. The majority of positive patients are from Europe and share the recessive AAGA founder risk haplotype (rs2066790, rs11096992, rs17584703, and rs6844176), which spans *RFC1* gene. *RFC1* gene encodes replication factor complex subunit 1 which makes a complex with other four subunits of replication factor complex, RFC. RFC helps in loading of PCNA onto DNA templates and thus helping in replication and repair by DNA polymerase ο and χ^15,16^. Altogether, biallelic (AAGGG)_exp_ at this locus has been found as causal mutation in neurodegenerative movement disorders like cerebellar ataxia, neuropathy, and vestibular areflexia syndrome (CANVAS), Late-onset ataxia (LOCA), Chronic idiopathic axonal polyneuropathy (CIAP) cases, Parkinson’s cases ^17,18, 19–26, 27–29^. The most common symptom since disease onset is progressive unsteadiness in *RFC1* TNR expansion positive cases.

In the present study, we investigated the prevalence of biallelic AAGGG expansion in Indian ataxia and other neurological disorders, variability of repeat motif and repeat size at *RFC1* repeat locus in Indian population-based disease cohorts and haplotype analysis in Indian background population. In literature, this expansion mutation does explain ~91% of sporadic CANVAS cases, ~100% familial CANVAS cases, >30% cases of hereditary neuropathy, >20% cases of late onset ataxia cases and also found in Parkinson’s disease cases. Since tandem repeat expansion in *RFC1* gene has been linked to a wide range of symptoms, there are some overlapping symptoms between CANVAS and SCA and CMT. Therefore, we chose to study cases from SCA, and CMT cohorts.

## Materials and methods

### Patient inclusion and sample collection

We performed this study on three genetically uncharacterized Indian population-based neurological disease cohorts, i.e., uncharacterized ataxia, Spinocerebellar ataxia 12 (SCA12) and Charcot-Marie-Tooth (CMT) disease and healthy controls. Ethical clearance for this study was obtained from the Institutional Human Ethics Committee of CSIR-IGIB (IHEC). First, we included all index patients with ataxia who were evaluated at the ataxia clinic in AIIMS, Delhi, India and fulfilled the following criteria: (1) ataxia as a prominent clinical feature; (2) All cases had previously tested negative for spinocerebellar ataxias (SCA), SCA1, SCA2, SCA3, SCA6, SCA7, SCA10, SCA12, SCA17 and Friedreich ataxia (FRDA). Out of all the uncharacterized ataxia cases, 87 ataxia cases who were clinically suspected for SCA12 were taken as a separate group during analyzing the allelic and genotypic distribution of different repeat sizes. Second, Charcot-Marie-Tooth (CMT) disease or HMSN cases screened in this study were negative for *PMP22* gene duplications or deletion. We included healthy controls who do not have any neurological disorders. Their DNA was isolated by salting-out method or Lucigen’s genomic DNA isolation kit. Next, IndiGen population genome sequencing data was used for the estimation of the repeat variability, repeat type configuration and associated risk haplotype in background Indian population (with 1029 healthy individuals).

### Patient consent and confidentiality

Signed consent has been collected from all the patients included in this study. Suspected cases have been given IDs (I1-I32) while maintaining the patient privacy. There is additional set of IDs (RFC1-8892 and RFC1-108) mentioned for few samples which are solely created for research purpose and do not disclose any information of patient identity. We have reported only those personal and clinical information of patients which are essential for understanding and interpreting the disease and causal mutations. We did not use patient’s identifiable information anywhere in the manuscript.

### Genetic screening of AAAAG locus at *RFC1*

We adopted two strategies to screen DNA samples of 1818 uncharacterized ataxia samples (females: 543, Avg. Age: 36-40 yrs), 93 clinical CMT (or HMSN) cases (females: 32, Avg. Age: 21-25 yrs), 87 ataxia samples (with SCA12 clinical diagnosis; Gender and Age: NA) and 564 controls (female: 268, Avg. age: 31-35 yrs) for *RFC1* TNR locus as shown in Figure 1a. Out of 1818 ataxia samples, 773 samples were directly undergone RP-PCR first for ‘AAGGG’ repeat motif and those with ‘AAGGG’ expansion were checked further for other two repeat motifs’ expansion (i.e., AAAGG and AAAAG). While, for rest of the cases and controls, we performed short-range flanking PCR (f-PCR) and repeat-primed PCR (RP-PCR) for AAAAG, AAAGG and AAGGG as described in Figure 1. Primers were used as given in the article by Cortese et al, 2019. Capillary electrophoresis was performed using ABI 3730xl DNA Analyzer and ABI 3500 Genetic Analyzer and data analysis was done using GeneMapper software.

**Figure 1:**
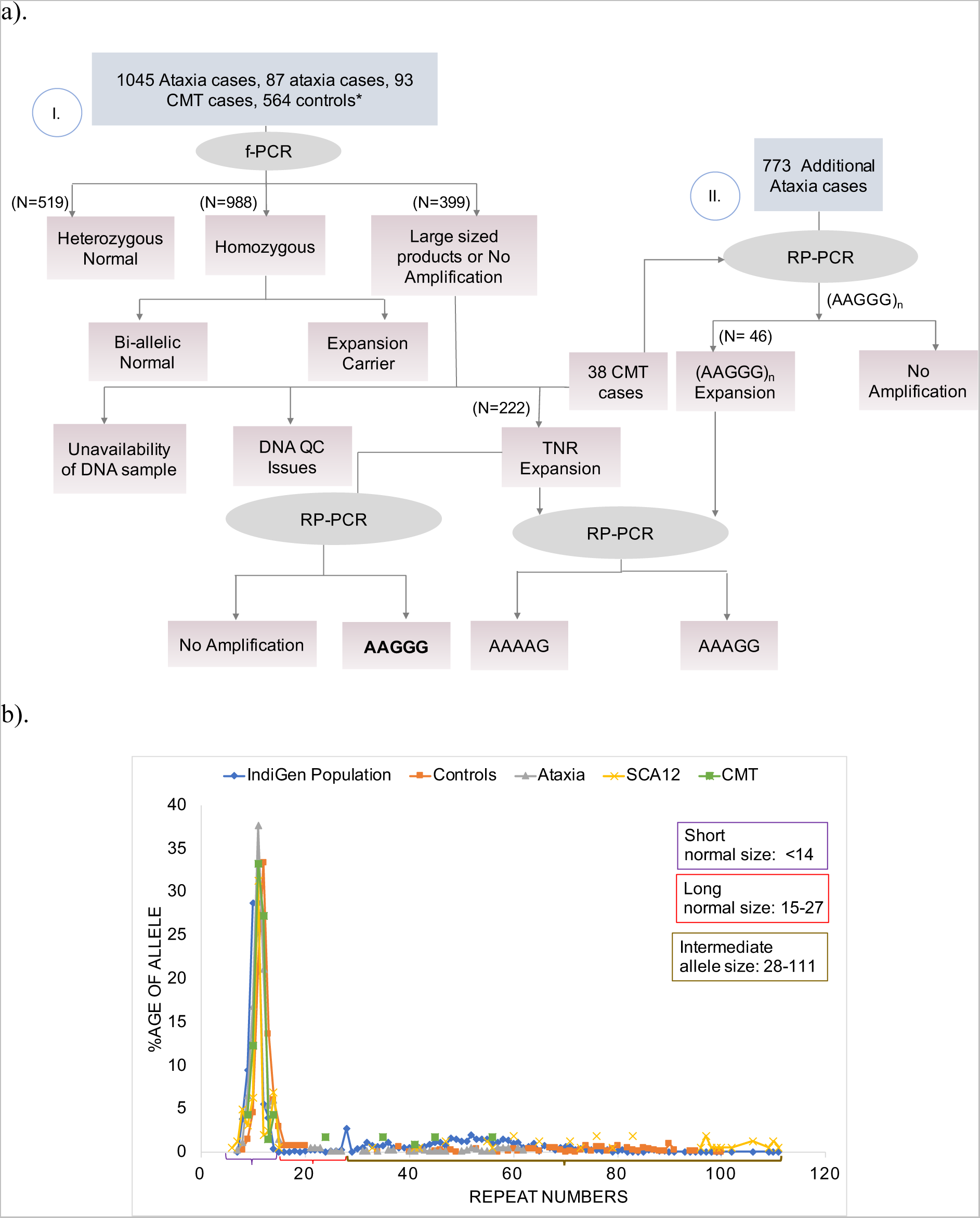
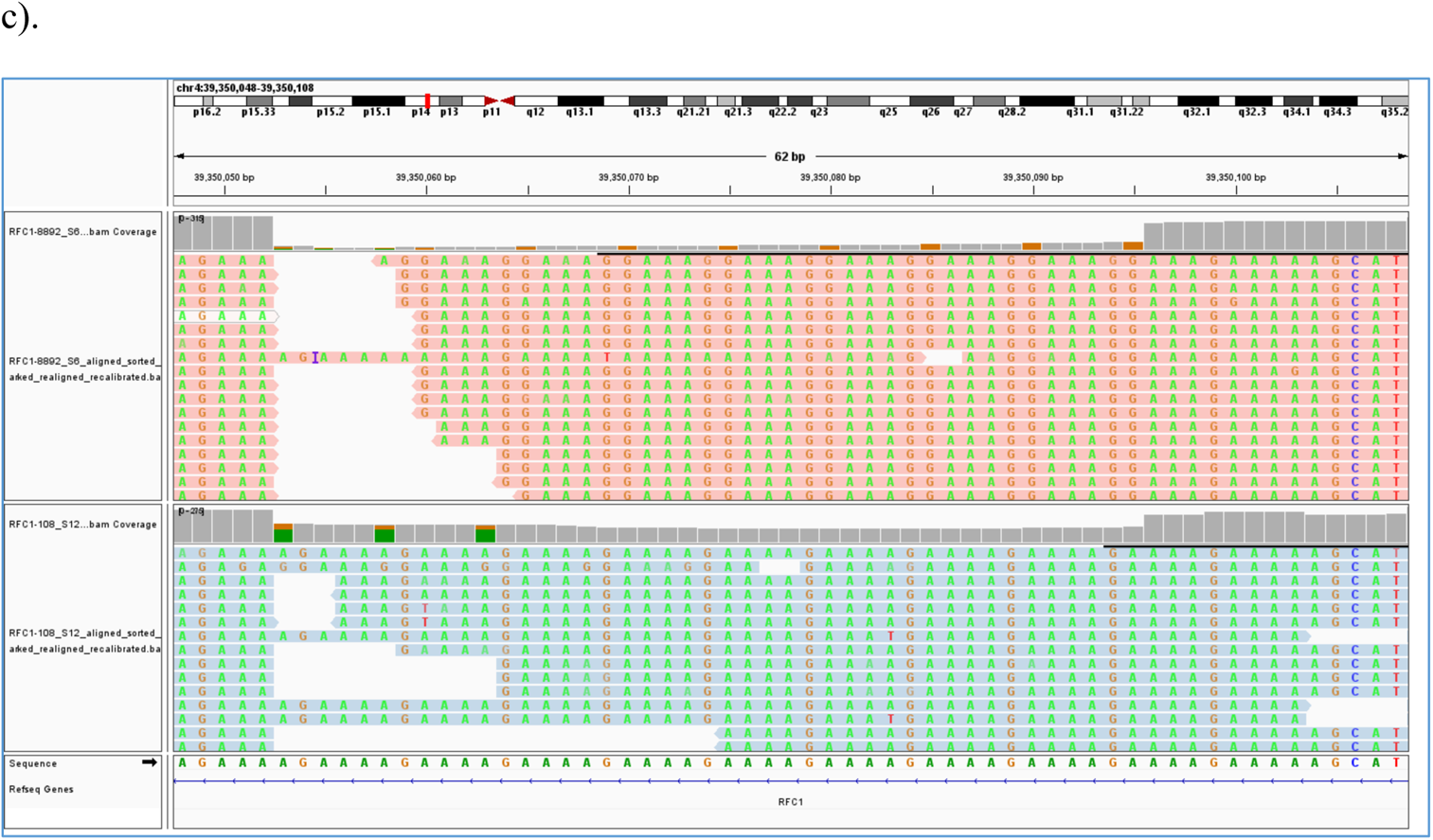
Genetic screening at RFC1 locus in Indian cohorts. a). Strategy to screen cases and controls for TNR locus at RFC1 gene in Indian context. (*We performed both f-PCR and RP-PCR in all the heterogenous controls) b). Allele frequency distribution at *RFC1* repeat locus in IndiGen population (886), controls (460), ataxia (809), SCA12 (80) and CMT (57) groups. (Here, total number of cases corresponding to each studied group represents those whose *RFC1*-TNR size was estimated using flanking PCR and are normal) c). miSeq sequencing of (AAAGG)_exp_ allele in (AAAGG)_exp_/(AAGGG)_exp_ positive case, I24 (RFC1-8892) and of (AAAAG)_exp_ allele in a (AAAAG)_exp_/(AAAGG)_exp_/(AAGGG)_exp_ carrier, I19 (RFC1-108).

Primarily, we performed f-PCR to find the cases with pathogenic expansion at *RFC1* locus. Additionally, we gathered f-PCR repeat size data for all the subjects studied in order to better understand the risk by repeat size or association of repeat size with disease group. Our f-PCR analysis was limited to estimate repeat size till 111 repeat numbers (Figure 1b).

Long range PCR was performed using primers (FP: 5’ GACATTACCATTCCAAAGAGGAG 3’, RP: 5’ CCTGAGTCCTCCTGACTGCT 3’) to amplify the whole TNR locus in *RFC1* gene in suspected RP-PCR positive and carrier cases (using LA-taq HS polymerase with Lucigen’s FailSafe G). PCR cyclic conditions were 98°C for 4’, 18 cycles of (98°C for 30”, 65°C for 15”, 72°C for 3’) with decrease of 0.5°C per cycle, 18 cycles of (98°C for 30”, 58°C for 15”, 72°C for 3’), 72°C for 10’. NGS library was prepared with amplified PCR products using Nextera XT, sequenced on Illumina’s MiSeq platform and analyzed using ExpansionHunter denovo (Supplementary Figure S1). Sanger sequencing was done on selected PCR amplified samples to validate RP-PCR results. (including both patients and control samples) (Supplementary Figure S5). Suspected samples, both carriers and biallelic (AAGGG)_exp_ cases were genotyped for the ‘AAGA’ risk haplotype using SNaPshot and were confirmed by Sanger sequencing. Primers used in this experiment are listed in Supplementary Table 5.

### Analysis of AAAAG TNR locus in Indian background population

Repeat numbers and different repeat motifs at *RFC1* locus were estimated in high throughput data of IndiGen population^30^ using Expansion Hunter and ExpansionHunter Denovo respectively ^31,32^. This population has a total of 1029 heathy Indian individuals though we could retrieve repeat number and repeat motif data for 886 and 678 individuals respectively. We compared genotypic frequency distributions across different repeat size categories between IndiGen population and other groups (Ataxia, SCA12, CMT and controls). Significance of association between repeat size-based genotype categories and repeat types (motifs or patterns) was determined using χ^2^ test with or without Yates’s correction.

We also retrieved genotype data of target SNPs (risk haplotype), from whole genome sequencing data of IndiGen population^30^. We aligned the individual-wise genotype data with repeat motif data and repeat number data separately. Haplotypes were phased along with repeat number and repeat type using PHASE software as two different analysis^33^. Next, with the phased data we attempted to interpret the relationship between *RFC1* TNR repeat number and associated haplotype and also between *RFC1* TNR locus repeat motif and associated haplotype using PopART. We had both haplotype and repeat number data for 886 healthy individuals. Additionally, we had haplotype and repeat type data for 678 individuals. Thus, we proceeded our analysis with 678 individuals for whom we have all the three data types. For this purpose, first, we categorized the repeat numbers into three repeat size classes: small normal (repeat numbers: 7-14, total allele count: 923); long normal (repeat numbers: 15-27, total allele count: 44), and intermediate length (repeat numbers: 28-111, total allele count: 805) and we gave them coding like S, L, and I respectively. Frequency of different haplotypes (count and percentage) in IndiGen population is given in Table 6. We transformed this data into trait file format and haplotype data into alignment file format as required in PopART software. Similarly, second analysis was performed for repeat motif and haplotype data. To note, repeat categories in the phylogenetic study are different from those in genotype-disease association study or genotype-repeat type association study because of the representation limitation in PopART.

### Data availability

miSeq data of suspected patients is not provided with the manuscript and will be shared on the request to corresponding author.

## Results

### Screening of known pathogenic A_n_G_n_ configuration in patients of Ataxia and CMT

Repeat size were determined from the f-PCR data for each group except 773 ataxia samples which underwent the alternate screening strategy as given in Figure 1a. Of all ataxia samples screened by f-PCR and repeat primed PCR (RP-PCR) (Supplementary Table 1 & Figure 1), three patients had biallelic (AAGGG)_exp_ expansions as defined by no amplifiable product in f-PCR, and a positive RP-PCR for AAGGG motif expansion. There was a total of thirty-two suspected ataxia cases with AAGGG TNR expansion in at least one allele, accompanied by either expansion of AAAGG or AAAAG or both AAAGG and AAAAG. In SCA cohorts, a total of six cases with (AAAGG)_exp_/(AAGGG)_exp_ and seven cases with (AAAAG)_exp_/(AAAGG)_exp_/(AAGGG)_exp_ were found with saw-tooth decremental pattern specifically in (AAGGG)_exp_. Out of clinical CMT samples, two cases with only AAGGG expansion, three cases with (AAAGG)_exp_/(AAGGG)_exp_ and four cases with (AAAAG)_exp_/(AAAGG)_exp_/(AAGGG)_exp_ were found. Out of screened control samples, we found three individuals with (AAAGG)_exp_/(AAGGG)_exp_ TNR expansion profile with AAGGG motif only with <50 repeat numbers (Table 1). Long range PCR (LR-PCR) was performed in thirty-two suspected ataxia cases (namely, I1 to I32) and nine suspected CMT cases. Long range PCR could amplify a largest product of around 4.5kb size which corresponds to ~700 repeats in one of the suspected ataxia cases.

**Table 1:** Count and percentage of cases and controls with repeat expansion (RP-PCR results) (cells with bold text have higher percentage than in controls). *Controls having repeat expansion within normal range. (‘n’ represents total number of cases in a particular group)

|  | (Strategy I) |  |  |  | (Strategy II) |
| --- | --- | --- | --- | --- | --- |
| Repeat type<br>(Expansion) | Controls<br>(n=564) | Ataxia<br>(n=1045) | CMT<br>(n=93) | SCA12<br>(n=87) | Ataxia (n=773) |
| AAAGG/AAGGG | 3*<br>(0.53%) | 1 (0.1%) | 3<br>(3.23%) | 0 (0%) | 5 (0.65%) |
| AAGGG/AAGGG | 0 (0%) | 0 (0%) | 2<br>(2.15%) | 1 (1.15%) | 2 (0.26%) |
| AAAAG/AAAAG | 4<br>(0.71%) | 1 (0.1%) | 0 | 4 (4.5%) | NA |
| **AAAAG/AAAGG** | 1<br>(0.18%) | 2 (0.19%) | 0 | 4 (4.5%) | NA |
| **AAAAG/AAGGG** | 1<br>(0.18%) | 2 (0.19%) | 0 (0%) | 0 (0%) | 7 (0.91%) |
| **AAAGG/AAAGG** | 10<br>(1.77%) | 1 (0.1%) | 0 | 0 (0%) | NA |
| **AAAAG/AAAGG/A**<br>**AGGG** | 0 (0%) | 2 (0.19%) | 4 (4.3%) | 0 (0%) | 5 (0.65%) |

When amplified samples were sequenced using Illumina miSeq platform, seventeen samples were properly sequenced and analyzed using EHdn (ExpansionHunter denovo). To note, they exhibited additional repeat motifs like AAAGGG, AAGAG, and AAGGC other than AAAAG and AAAGG whereas AAGGG motif was surprisingly missing (Supplementary Figure S4). One reason could be the G-rich nature of AAGGG TNR expanded DNA sequence which might have hindered the amplification of allele containing AAGGG repeat motif expansion in long range PCR. Profiles like, AAAGG, AAAAG, AAAGGG and AAGAG were visualized using Sanger sequencing also. AAAGGG, a hexanucleotide repeat motif was found in 4/17 carriers (Supplementary Figure 1).

Out of three SCA patients with biallelic (AAGGG)_exp_ in *RFC1* gene, only one case (I32) had recessive ‘AAGA’ haplotype and had symptoms similar to CANVAS (Table 2). Additionally, three out of six (AAAGG)_exp_/(AAGGG)_exp_ SCA cases also exhibited recessive risk haplotype. Of them, I24, a SCA case also presented phenotypes similar to CANVAS. AAAGG expansion profile of I24 has been shown in Figure 1c. No suspected case in CMT disease cohort had recessive risk haplotype.

**Table 2:** Clinical details of RFC1-TNR expansion positive patients. (NA: data not available)

| **Sample ID** | **I33** | **I24** | **I32** | **I14** | **I6** |
| --- | --- | --- | --- | --- | --- |
| **Age (Age of onset)** | 26-30<br>(21-25) | 51-55(36-40) | 56-60<br>(56-60) | 26-30 | 46-50 |
| **Sex** | Male | Female | Female | Male | Male |
| **Clinical Diagnosis** | SCA<br>PANEL | SCA PANEL | SCA | SCA | SCA |
| **SCA Panel Screening (negative)** | Essential Tremor | SCA1, SCA2, SCA3, SCA6, SCA12 and SCA17 | SCA1, SCA2, SCA3, SCA6, SCA12 and SCA17 | Autosomal dominant cerebellar ataxia | Autosomal dominant recessive ataxia, Autosomal recessive cerebellar ataxia |
| **RFC1-TNR profile** | (AAAGG)<sub>x</sub>/(AAGG G)<sub>exp</sub> | (AAAGG)<sub>x</sub>/(AAGGG)<sub>exp</sub> | (AAGGG)<sub>exp</sub>/(AAGGG)<sub>exp</sub> | (AAAGG)<sub>x</sub>/(AAGGG)<sub>exp</sub> | (AAAAG)<sub>x</sub>/(AAGGG)<sub>exp</sub> |
| **Haplotype** | AAGA | AAGA | AAGA | AAGA | AA(G/A)(A/G) |
| **First symptom** | Tremor in both hands (postural and intentional<br>) | Imbalance while standing, weakness | Walking difficulty | NA | NA |
| **Inheritance** | Sporadic | Sporadic | Sporadic | NA | NA |
| **Past history** | NA | Cough since<br>childhood | Coughing while<br>eating (10yrs<br>ago) | NA | NA |
| **Family**<br>**descendants** | NA | NA | Mentally<br>retarded child,<br>cannot speak | NA | NA |
| **Neurological**<br>**condition** | Head<br>tremor,<br>hand<br>tremor,<br>speech<br>problem | Gait ataxia,<br>writing<br>problem,<br>dysphagia | Gait Ataxia,<br>head tremor,<br>speech problem,<br>dysdiadokokine<br>sia, nystagmus,<br>broken pursuit | NA | NA |
| **Other**<br>**medical**<br>**condition** | NA | Thyroidism,<br>muscle<br>cramps,<br>vision<br>problem,<br>muscle<br>cramps,<br>reduced sleep | Vitamin E<br>deficiency | NA | NA |
| **Nerve**<br>**conduction**<br>**test** | NA | Small fiber<br>neuropathy | Vibrations<br>reduced in UL<br>and LL | NA | NA |
| **Sympathetic**<br>**reactivity** | NA | Mild loss | NA | NA | NA |
| **Cardiac**<br>**Autonomic**<br>**tone** | NA | Severe loss | NA | NA | NA |

### Case Studies

Clinical data of expansion positive cases has been shown in Table 2.

I32 (Biallelic-(AAGGG)_exp_)

Patient is a female and was SCA1, 2, 3, 6, 12 and SCA17 negative. She had slowly progressive difficulty in walking at the time of reporting. Two years before the day of reporting, patient presented only the issue of cough while eating and since then condition progressed such that, later on, patient required the ADL support.

I24 ((AAAGG)_exp_/(AAGGG)_exp_)

This patient is female who had imbalance while walking and patient have cough since childhood. Patient is SCA1, 2, 3, 6, 12, SCA17 and FRDA negative. Majorly, this patient has small fiber neuropathy, cerebellar ataxia, imbalance while standing and loss of cardiac autonomic tone. Cough was present in paternal family also.

### Association of disease group and repeat type with the tandem nucleotide repeat size

Frequency distribution of repeat numbers vs allele frequency in different disease groups and controls is plotted (Table 3). Strangely, in all the cohorts (control and case group) studied here, repeat sizes, 15-27 were rare in all the study groups and thus creating a clear break/drop in allele frequency distribution (Figure 1b & Table 3 & table 4). In IndiGen repeat size distribution, a striking drop in allele frequency is observed from repeat size 14 to 15, while again there is increase in allele count beyond repeat size, 27. Based on these observations from repeat size distributions at *RFC1* locus in IndiGen healthy population and other cohorts (as shown in Figure 2a), repeat lengths were categorised into three repeat size bins, SN-small normal (repeat numbers: 7-14); LN-long normal (repeat numbers: 15-27), and IA-intermediate length allele (repeat numbers: 28-111) in order to calculate a more precise risk for disease in relation to *RFC1*-TNR length. Allelic distribution of different groups based on repeat size categories is shown in Table 3. Additionally, repeat range, SN was found significantly associated with other groups analysed in lab but IndiGen population (Supplementary Table 2). On the contrary, we found repeat range, IA associated with IndiGen population. This contrasting difference between IndiGen population and other studied groups in lab could be due to different approaches used to generate repeat size data. Surprisingly, allele frequency of repeat category, “long normal”, is significantly lower in ataxia group.

**Figure 2:**
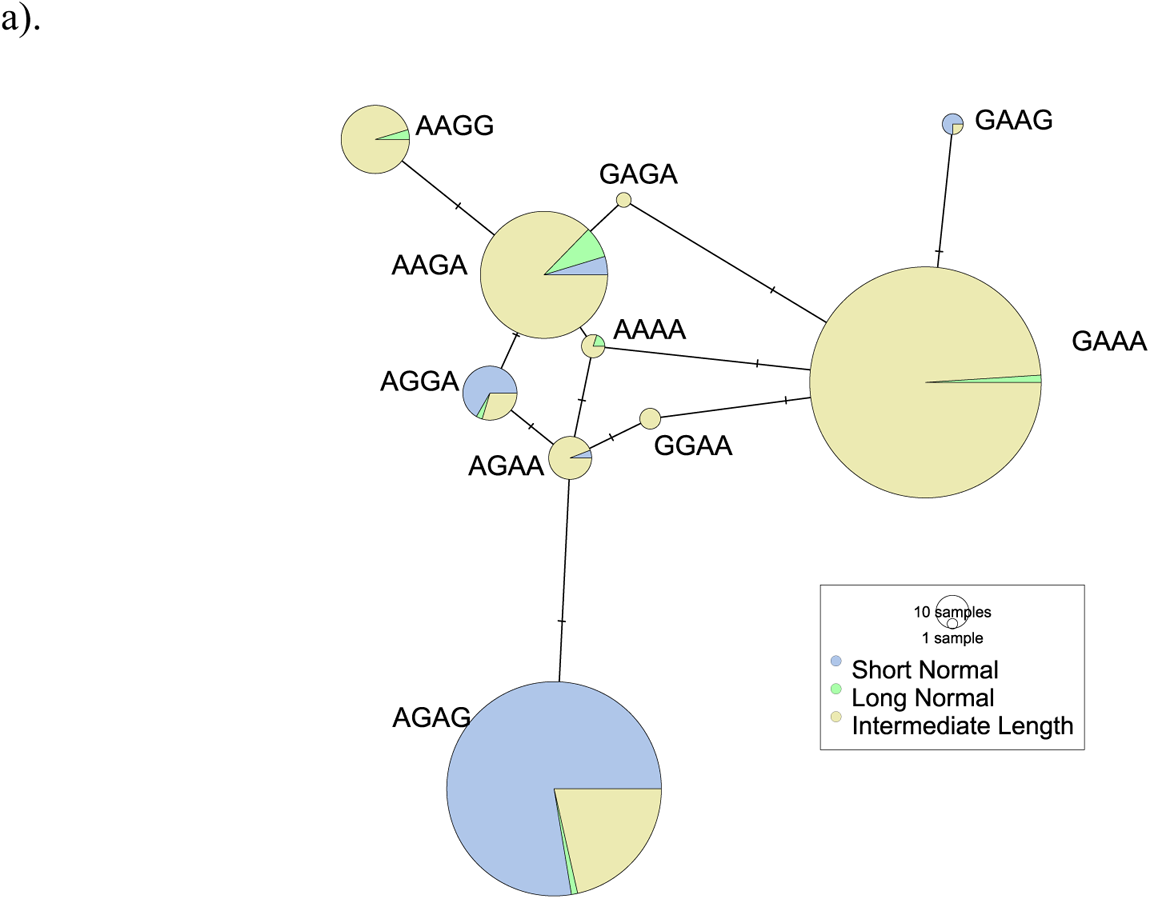
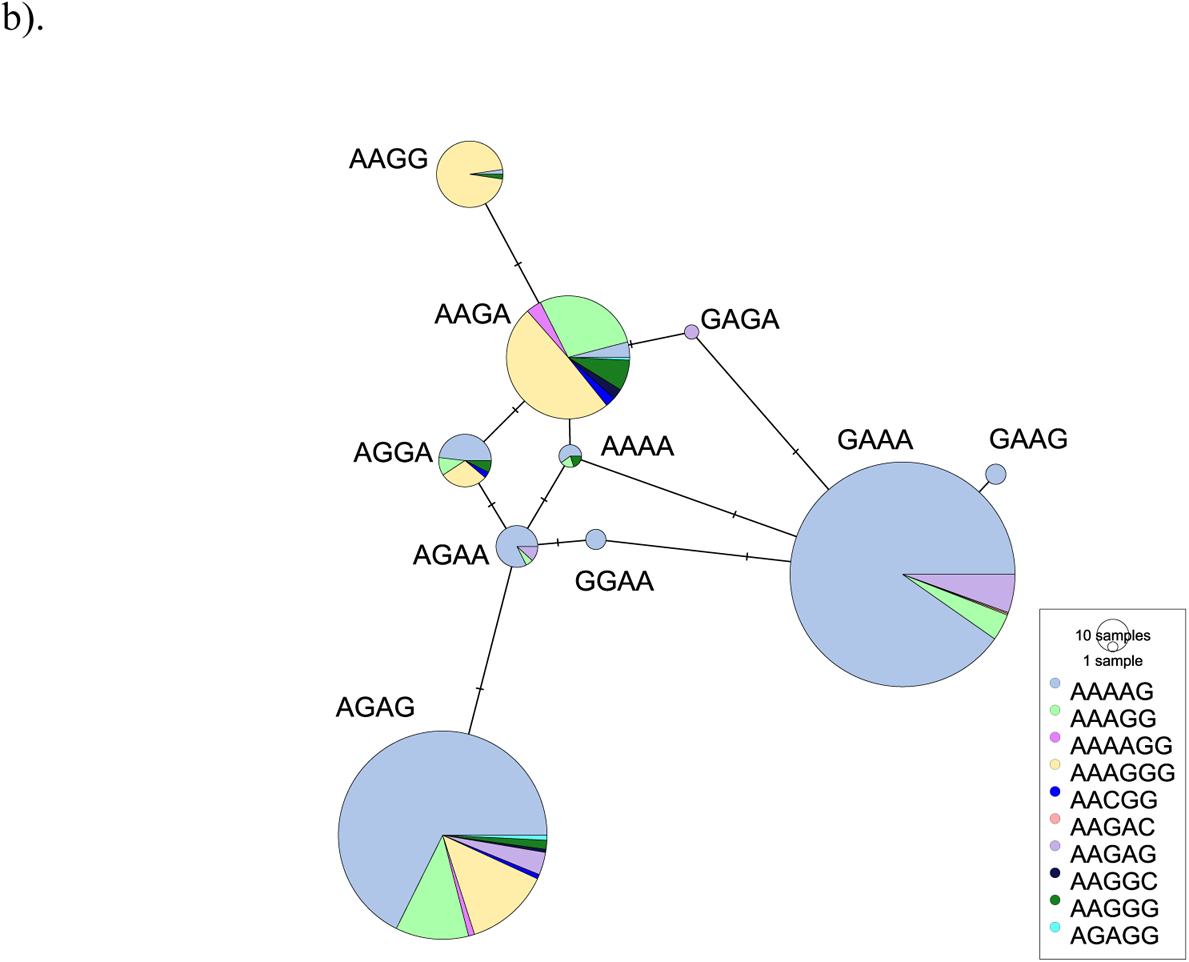
Phylogenetic representation of relationship between haplotype and repeat size in IndiGen population: Median spanning phylogenetic network of individual-wise target haplotype and RFC1-TNR number. b). Phylogenetic representation of relationship between haplotype, repeat type: Median spanning phylogenetic network of individual-wise target haplotype and RFC1-TNR type. (Short normal: <14; Long normal: 15-27; Intermediate allele: 28-111)

**Table 3:**
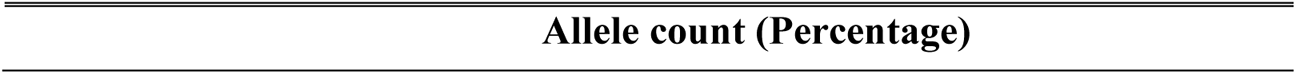
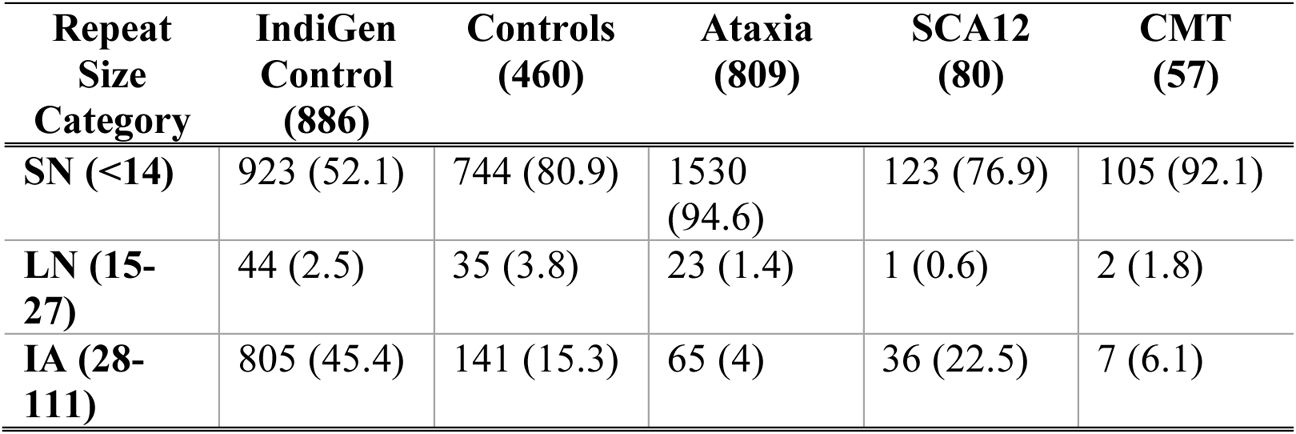
Allele distribution across different repeat size range in different study groups. (Here, number of cases corresponding to each study group represents those whose *RFC1*-TNR size could be estimated using flanking PCR and are normal)

**Allele count (Percentage)**
| **Repeat**<br>**Size**<br>**Category** | **IndiGen**<br>**Control**<br>**(886)** | **Controls**<br>**(460)** | **Ataxia**<br>**(809)** | **SCA12**<br>**(80)** | **CMT**<br>**(57)** |
| --- | --- | --- | --- | --- | --- |
| **SN (<14)** | 923 (52.1) | 744 (80.9) | 1530<br>(94.6) | 123 (76.9) | 105 (92.1) |
| **LN (15-**<br>**27)** | 44 (2.5) | 35 (3.8) | 23 (1.4) | 1 (0.6) | 2 (1.8) |
| **IA (28-**<br>**111)** | 805 (45.4) | 141 (15.3) | 65 (4) | 36 (22.5) | 7 (6.1) |

**Table 4:** Repeat size-based genotype distribution at *RFC1* locus in different cohorts. (Here, number of cases corresponding to each study group represents those whose *RFC1*-TNR size could be estimated using flanking PCR and are normal)

| **Genotype count (Percentage)** |  |  |  |  |  |
| --- | --- | --- | --- | --- | --- |
| **Genotypes** | **IndiGen**<br>**Control**<br>**(886)** | **Controls**<br>**(460)** | **Ataxia**<br>**(809)** | **SCA12**<br>**(80)** | **CMT**<br>**(57)** |
| SN\|SN | 285<br>(32.2) | 336 (73) | 751<br>(92.8) | 58 (72.5) | 51 (89.5) |
| SN\|LN | 24 (2.7) | 15 (3.3) | 9 (1.1) | 1 (1.3) | 0 (0) |
| SN\|IA | 329<br>(37.1) | 57 (12.4) | 19 (2.3) | 6 (7.5) | 3 (5.3) |
| LN\|IA | 8 (0.9) | 2 (0.4) | 0 (0) | 0 (0) | 0 (0) |
| IA\|IA | 234<br>(26.4) | 41 (8.9) | 23 (2.8) | 15 (18.8) | 2 (3.5) |
| LN\|LN | 6 (0.7) | 9 (2) | 7 (0.9) | 0 (0) | 1 (1.8) |

At genotype level, we compared IndiGen control population with all the other groups. A total of 6 genotypes were present with repeat categories (SN, LN and IA). In order to further analyse *RFC1* TNR locus between IndiGen control group and other study groups at genotype level, we estimated the genotype count based on repeat size categories in different studied groups. Genotypic distribution of IndiGen controls, heterogeneous lab controls, Ataxia, SCA12 and CMT groups has been shown in Table 4. We found that the count (percentage) of genotypes, SN|LN, LN|IA and LN|LN were very low in all the groups (IndiGen controls, controls, Ataxia, SCA12 and CMT). Whereas, SN/SN, which was homozygous small normal, was found significantly associated with all the study groups in comparison to IndiGen population (Supplementary Table 3). IndiGen controls might be showing different patterns than lab controls and other groups again because the repeat data has been generated through different approach in IndiGen population compared to other groups. To note, SN|LN and LN|IA were found significantly rare in ataxia group in comparison to IndiGen population.

In IndiGen population, we found nine distinct repeat motifs at *RFC1* repeat locus accounting for 678 individuals. Percentage of different genotypes in IndiGen population is given in Supplementary Figure 2. Briefly, apart from AAAAG repeat motif (biallelic-60% and rest-3%) and AAAGG (biallelic-5% and rest-8%), we found that 11% of IndiGen population exhibits biallelic hexanucleotide repeat motif, AAAGGG, while in 5% of IndiGen population, only one allele has AAAGGG repeat motif. 1.33% of IndiGen population exhibits biallelic AAGGG pattern. However, in IndiGen population, we were unable to locate any individuals with the ACAGG and AAAGG/AAGGG repeat motif arrangement.

Next, at genotype level, we compared different repeat motifs at *RFC1* repeat locus in IndiGen population with repeat motif AAAAG (which was considered as normal/reference repeat motif) to observe any association with repeat size-based genotypes. Data of repeat type-based genotype frequency distribution in IndiGen population is shown in (Table 5) and association analysis results are given in detail in Supplementary Table 4. Interestingly, we found that genotypes having long normal allele have association with AACGG, AAGGC, AAGAG and AAAGGG repeat motifs. IA|IA genotypes have maximum number of diverse repeat types showing that with the increase in repeat size, there increases the chances of the presence of different repeat types.

**Table 5:**
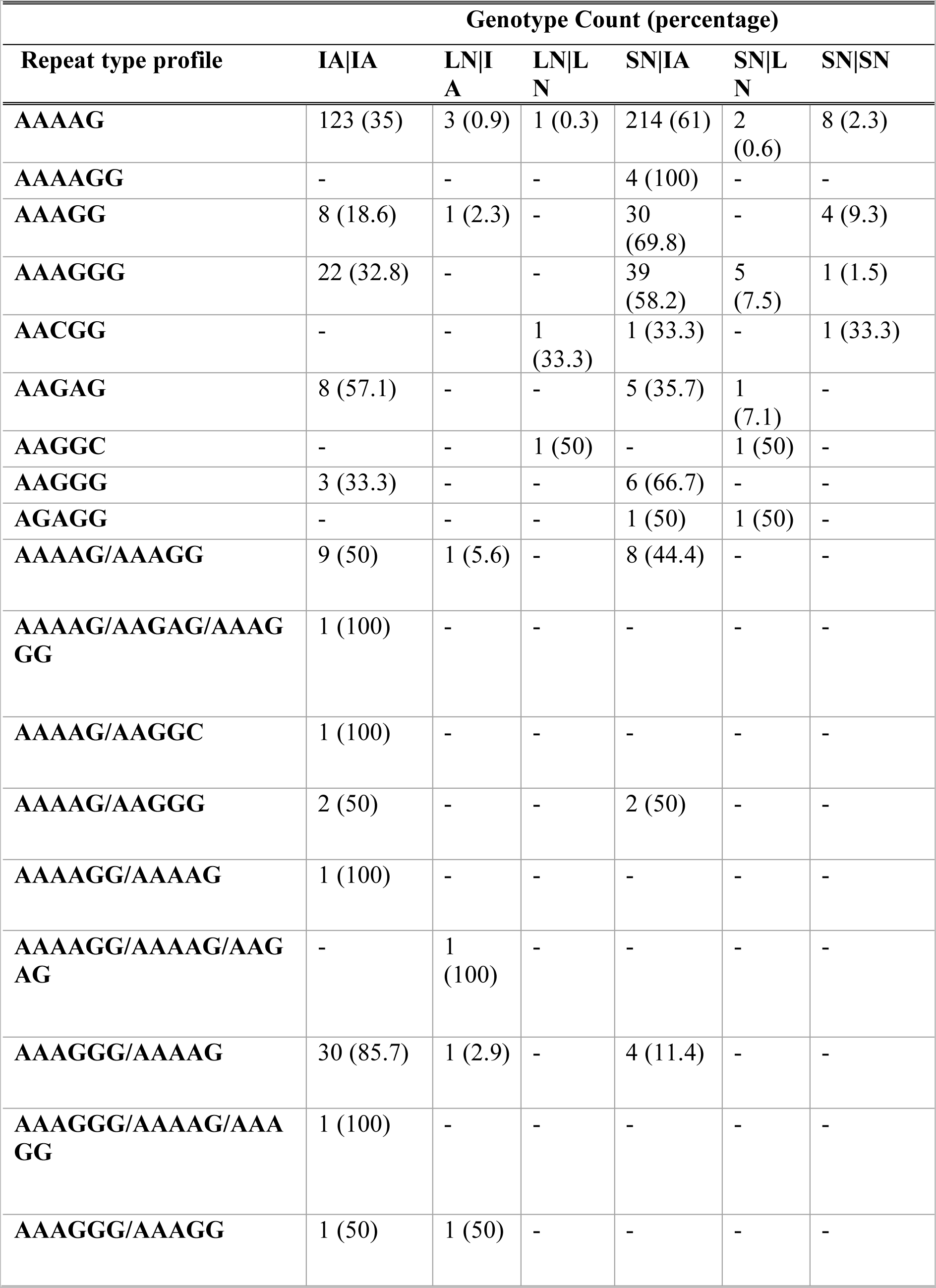
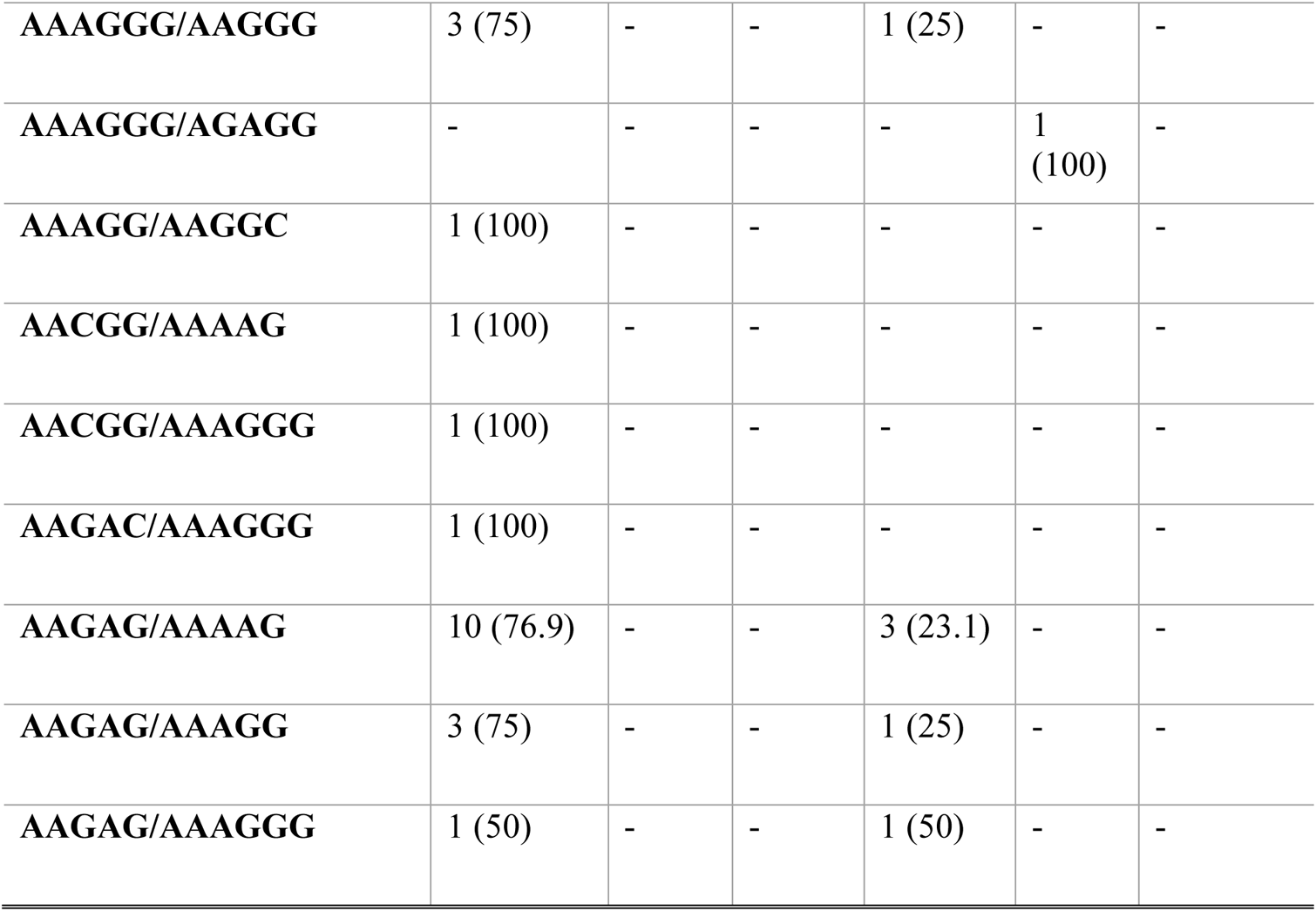
Genotype count (percentage) distribution between repeat number and repeat type in IndiGen population.

### Haplotype assessment in background population

The overall allele frequency of risk haplotype ‘AAGA’ was 8-9% in IndiGen population. Frequency of different haplotypes in IndiGen population distributed in three repeat size categories is given in Table 6. Phylogenetic representation shows that two haplotypes, ‘AGAG’ and ‘GAAA’ were prevalently present in IndiGen population (Figure 2). Individuals with ‘AGAG’ haplotype were diverse in repeat length and repeat types as well while individuals with ‘GAAA’ mostly exhibited intermediate repeat length and most prevalent repeat type AAAAG followed by AAGAG and AAAGG. However, this haplotype didn’t represent AAGGG repeat motif. On the other hand, third most frequent haplotype was risk haplotype, i.e., ‘AAGA’ in IndiGen population. Individuals with this haplotype exhibited diverse repeat types with most frequent AAAGGG repeat type. They exhibited all the three different repeat lengths with most frequent ‘intermediate length’ followed by long normal. 40.9% of long normal alleles exhibits ‘AAGA’ risk haplotype out of all the haplotypes. Surprisingly, two out of 1029 IndiGen controls had recessive risk haplotype ‘AAGA’ with repeat numbers, 30/32, 40/49 and repeat motifs, biallelic AAAGGG and AAAGG/AAAGGG respectively and thus are at-risk individuals.

**Table 6:** Frequency of different haplotypes (allele count and percentage) in IndiGen population (total allele count: 1772) in three different repeat size categories.

| **Repeat number**<br>**range →** | **7-14** | **15-27** | **28-100** |
| --- | --- | --- | --- |
| **Haplotypes** | **Small**<br>**Normal**<br>**(n=923)** | **Long**<br>**Normal**<br>**(n=44)** | **Intermediate**<br>**(n=805)** |
| AAAA | - | 1 (2.3%) | 4 (0.5%) |
| AAGA | 8 (0.9%) | 18 (40.9%) | 138 (17.1%) |
| AAGG | - | 2 (4.5%) | 41 (5.1%) |
| AGAG | 844 (91.4%) | 10 (22.7%) | 97 (12%) |
| AGGA | 60 (6.5%) | 4 (9.1%) | 9 (1.1%) |
| GAAA | - | 9 (20.5%) | 493 (61.2%) |
| AGAA | 4 (0.4%) | 16 (2%) | 4 (0.4%) |
| GAAG | 5 (0.5%) | - | 1 (0.1%) |
| GAGA | - | - | 2 (0.2%) |
| GGAA | - | - | 4 (0.5%) |
| GGAG | 2 (0.2%) | - | 2 (0.2%) |

## Discussion

According to our knowledge, our work is the first to study *RFC1* repeat locus in Indian population. Role of alleles with repeat types, (AAGGG)_<400_, (AAAGG)_<970_ and (AAAAG)_<1000_ and other rare repeat types have not yet demonstrated in disease risk or are otherwise stated as non-pathogenic in literature. We put efforts to study the association of different repeat size bins with cases and also the association of repeat size-based genotype with particular repeat motif in IndiGen control population at *RFC1* repeat locus.

*RFC1* repeat locus is highly dynamic in terms of tandem nucleotide repeat motif composition and repeat size in Indian population. Repeat type screening of *RFC1-*TNR locus in uncharacterised cases (SCA12: 87; Uncharacterised ataxia: 1818, CMT: 93, controls: 564) showed that the frequency of samples with bi-allelic (AAGGG)_exp_ are 1.15%, <0.05%, 2.15% and 0% respectively. We report new pathogenic expansion of (AAAGG)_exp_/(AAGGG)_exp_ in *RFC1* in three uncharacterised ataxia cases with recessive risk haplotype.

We found a total of four ataxia cases with recessive risk ‘AAGA’ haplotype, of them three cases had (AAAGG)_exp_ /(AAGGG)_exp_ and one case had biallelic (AAGGG)_exp_. In CMT cohort, we did not find any positive case who also had recessive risk haplotype. We had detailed clinical information for two (I32 and I24) out of four suspected ataxia patients and both of them represented cough earlier than first symptom of walking or standing imbalance. In our study, one (AAAGG)_~700_ was discovered at *RFC1* repeat locus in an Indian woman (I24) in her 60s who had clinical diagnosis of SCA. One or more of the CANVAS phenotypes are found in this patient. Besides her, there is occurrence of cough in family members who otherwise have clinically normal health is what distinguishes them as peculiar. The 700-AAAGG allele shared the same haplotype, according to haplotype marker analysis, indicating the existence of a common ancestor. The enlarged AAGGG (repeat number data is not available), discovered in the same patient, also had the same haplotype. This study also represents one sporadic SCA patient (I32) who had a biallelic pathogenic expanded AAGGG at *RFC1* repeat locus, ‘AAGA’ risk haplotype and clinical symptoms near similar to reported RFC1-related symptoms. Child of this case (I32) is presented with mental retardation and speech problem. This case study further makes it important for intermediate *RFC1* repeat alleles to take into the account the instability rate in the children of unaffected parents who have intermediate alleles that could exhibit lower penetrance and meiotic instability.

Cases who were carriers for (AAGGG)_exp_ also found to have AAAGGG, AAGAG and AAGGC TNR motifs from their NGS data analysis. Repeat motif like AAGGG, AAAGGG, AAAGG, AAGGC, AACGG, AAAAGG, AAGAG, AGAGG, AAGAG and AAAAG were found in IndiGen population at *RFC1* TNR locus except ACAGG. As AAAGGG is present in 16% of IndiGen population and it has also been found in expansion carriers. This further points towards studying these motifs for its pathogenicity. Huntington’s disease, SCA17, SCA2, SCA10 and ALS are repeat expansion disorders for which reduced penetrance alleles have been reported^34–37^. Our study does not conclude that a particular repeat size range imposes genetic susceptibility of disease on cases or their next generation exhibiting them but it opens up a question to understand it more clearly for disease’s late onset or onset in next generation.

There is a significant association of SN|SN (small normal homozygous) genotype with controls, ataxia, SCA12 and CMT as compared to IndiGen population and the reason could be that in this study we considered all the individuals with single peak in f-PCR data analysis (controls, ataxia, SCA12 and CMT) as homozygous while some of them might actually be heterozygous and expansion carrier with one allele large enough not to get amplified in given conditions of f-PCR. Additionally, genotypes with 15-27 repeats (long normal) are rare in all the study groups. Strangely, in all the cohorts (controls and cases) studied here, alleles with repeat size 17-20 are not found in controls or any case group except in IndiGen controls (11/886) where they were found in heterozygous form which alarms to study the possible reason of its absence in other study groups and to search the same in other population backgrounds. SN|LN and LN|IA were found significantly associated with IndiGen controls when compared to ataxia group. When we looked into repeat types exhibited by LN in IndiGen population, we found that they are significantly associated with either repeat motif, AAAGGG, or rare repeat motifs, AACGG, AAGAG, AGAGG and AAGGC as compared to AAAAG repeat motif. These observations suggest that in contrary to NGS in IndiGen population, f-PCR used for amplification in ataxia group could not amplify AAAGGG repeat motifs due to its complex nucleotide composition and rest of the motifs which are themselves very rare even in IndiGen population. Though, no of individuals with later repeat types are rare and thus association inferences might not be powerful enough to state upon significance levels. ‘AAGA’ risk haplotype is mostly present in heterozygous form in IndiGen population except otherwise in two controls (0.1%). These two individuals out of 1029 IndiGen controls have repeat numbers 30/32, 40/49 and the biallelic AAAGGG and AAAGG/AAAGGG repeat motifs, respectively.

We found more positive cases with (AAAGG)_exp_/(AAGGG)_exp_ at *RFC1* repeat locus in SCA cohort than biallelic-(AAGGG)_exp_ suggesting that (AAAGG)_exp_/(AAGGG)_exp_ configuration testing must be recommended for Indian SCA cases with sensory neuropathy. We found that ataxia cases with long normal repeat numbers are significantly lower in comparison to IndiGen controls. These repeat numbers are associated with repeat types like AAGAG, AGAGG, AACGG, AAGGC and AAAGGG which are GC rich motifs. Less number of such cases in ataxia group suggests that these motifs might have undergone expansion and additional RP-PCR assays for such motifs must be performed to rule out this hypothesis. Moreover, this study clearly marks the need of detailed clinical data of ataxia cases for recommendation of *RFC1* genetic testing. The presence of sensory neuropathy and vestibular areflexia along with cough in patients with cerebellar ataxia points towards the need of genetics testing of *RFC1* TNR locus.

The information from this study is important for making decisions about linking clinical interpretation to genetic research. Neuropathy is being accompanied by chronic cough in CANVAS and as cough precedes the first ataxic symptom, must be reported in clinical examination of patients for best decision making in further investigation into genetics. We advise clinicians to add CANVAS to their clinical interpretations for better and fast disease diagnosis. Even though cerebellar ataxia, dysautonomia, and cough can be absent, the diagnosis of CANVAS must be based on a profound vestibular deficit and axonal sensory neuropathy. This will further help to find the prevalence of CANVAS in Indian population and thus the *RFC1* repeat expansions on granular levels. This locus needs to be checked in other neurodegenerative disease cohorts with symptoms similar to CANVAS. With genetic heterogeneity at *RFC1* TNR locus comes wide spectrum of clinical symptoms and complex mode of pathogenesis at molecular level.

DNA samples for some suspected cases were not sufficient to perform LR-PCR and haplotype assessment and the further study was proceeded without them. LR-PCR on expanded repeat locus with high GC content could not be successful and requirement of enough sample for long read sequencing (we could not estimate exact repeat size) was not fulfilled. We could not be able to gather detailed clinical data for some cases positive for *RFC1* expansion which makes the diagnosis incomplete and indecisive for those cases. We still have a large number of uncharacterized cases which did not have any of the three repeat motifs (AAAAG, AAAGG and AAGGG) in the *RFC1* TNR locus screening. Exploring the genetic heterogenicity in current study in background population suggests to explore other rare repeat types in more uncharacterized cases in India.

The TNR loci that have longest pure tracts of repeats are expected to be among the most unstable and represent potential disease association prospects. So, it is necessary to have information of interruptions and repeat purity at *RFC1* locus. Long read sequencing techniques are to be adopted which are able to sequence longer DNA fragments without the need of PCR giving precise information at single nucleotide base level. This will further aid in better understanding how clinical heterogeneity is associated with genetic heterogeneity in *RFC1* related repeat expansions. It will help set the threshold for pathogenicity of repeat expansion which will help researchers working across the globe in tandem repeat expansion disorders.

## Data Availability

All the data will be provided upon request to corresponding author

## Abbreviations

CANVAS: cerebellar ataxia, neuropathy and vestibular areflexia syndrome
f-PCR: Flanking PCR
RP-PCR: Repeat Primed PCR
LR-PCR: Long range PCR
EHDn: ExpansionHunter denovo
IndiGen: Indian Genomes
ADL: Activities of Daily Living
STR: Short tandem repeats
TNR: Tandem nucleotide repeats

## Acknowledgement

We acknowledge Prof. Mitali Mukerji in initial direction towards this work. We sincerely appreciate everyone who has been constant support during this study including lab assistants, Subhash Gurjar, Usha Rawat and Suman Mudila. We are thankful for the participation of the patients and their families.

