## Supplementary File for "Investigation of *RFC1* tandem nucleotide repeat locus in diverse neurodegenerative outcomes in an Indian cohort"

**Supplementary figures**

**Supplementary tables**

Figure 1: Heat map showing the presence of different repeat units find in RFC1 in (AAGGG)_exp_ carriers

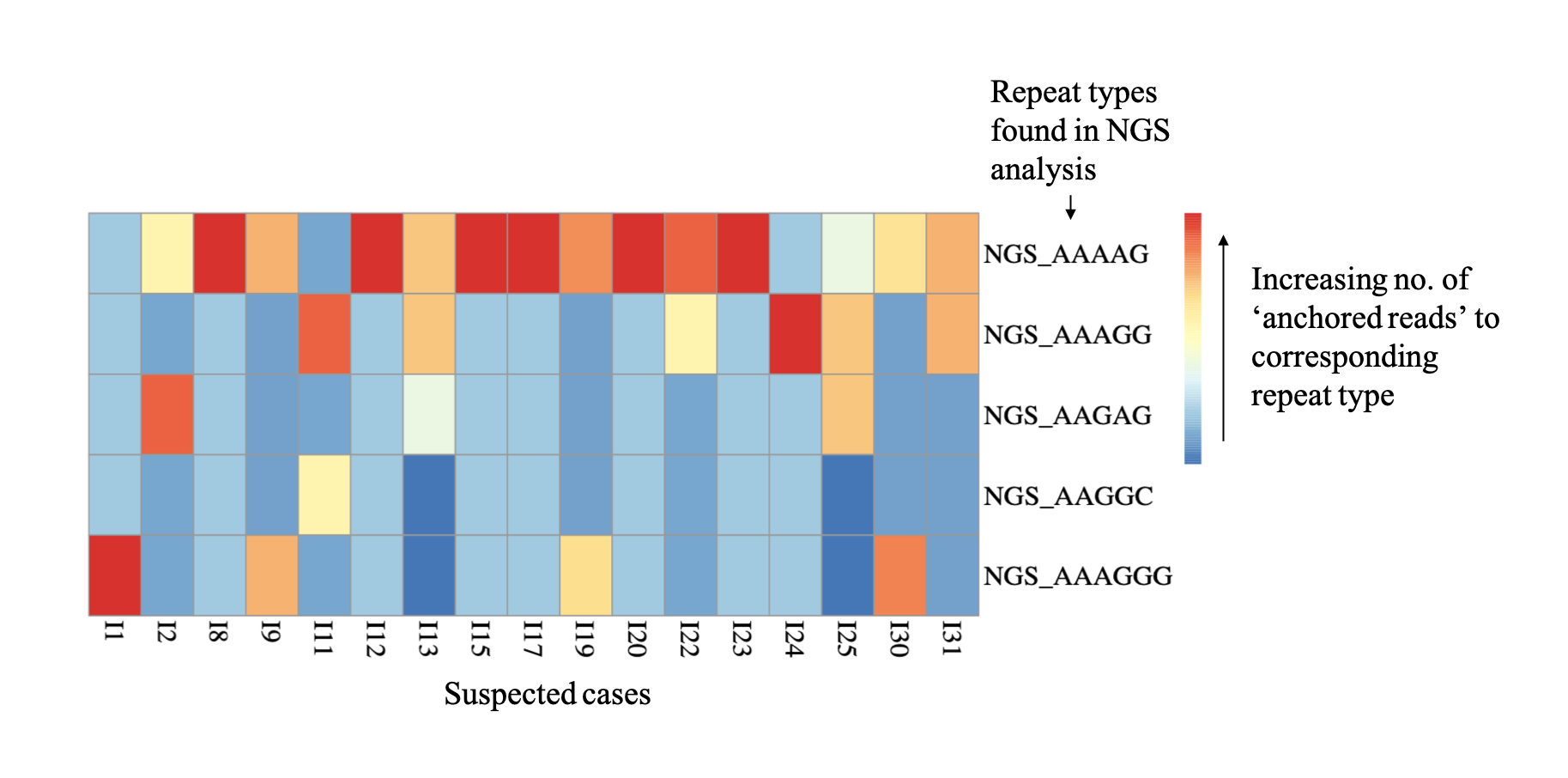

Figure 2: RFC1-TNR locus heterogeneity in IndiGen population (N=678).

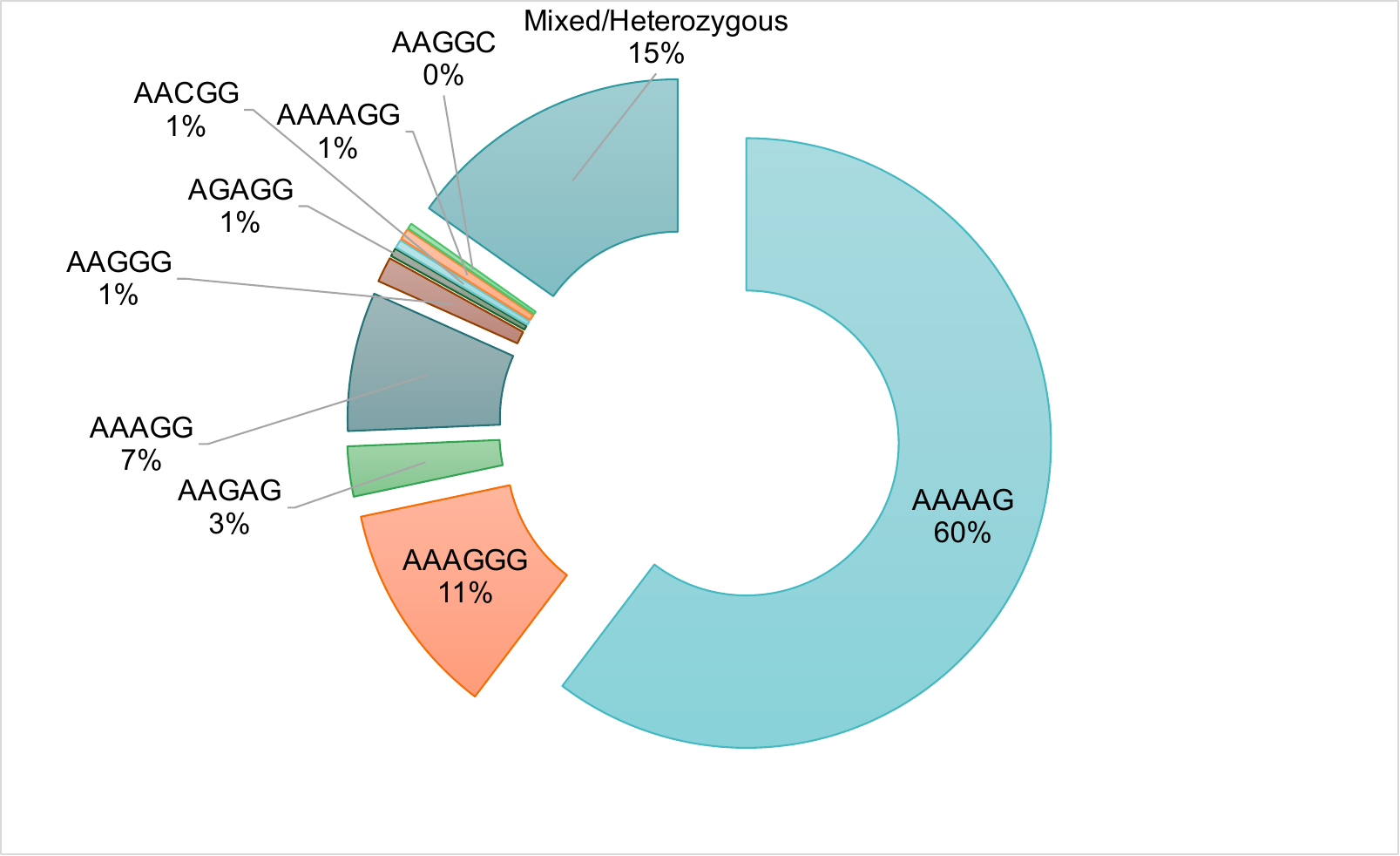

Table 1: Investigation of RFC1 locus in different groups using flanking PCR and RP-PCR. (#Few samples were not analysed in RP-PCR due to sample quantity limitation.)

a. Heterogenous controls

|  | **Flanking-PCR** | **Number of individuals with repeat expansions (Repeat-Primed PCR)** | | |
| --- | --- | --- | --- | --- |
| **Total (n=564)** | **Length of repeats (No of individuals)** | **AAAAG** | **AAAGG** | **AAGGG** |
| **Single Allele amplification (n=259)** | 8-16 (226), 41-91 (33) | 5 (A4G) | 16 (A3G2); 5 subjects with repeats 62, 69, 75, 80 and 89. 11 (A3G2/A2G3); one with 91 repeats | 8 (A2G3); 1 subject with 68 repeats, 1 subject with 64 repeats. |
| **Heterozygous (n=201)** | 9-16; 26 subjects with one allele having repeats >34 | - | 4(A3G2); 2 subjects with repeats 11/94 and 14/94. 1(A3G2/A2G3) | 14(A2G3); 3 subjects with 42/76, 12/91 and 11/99 repeats |
| **No amplification (? Homozygous expansion); n=104^#^** | - | 2(A4G) | 5(A3G2), 6(A3G2/A4G) | 3(A2G3);  3(A2G3/A3G2/A4G) |

b. Ataxia

|  | **Flanking-PCR** | **Number of individuals with repeat expansions (Repeat-Primed PCR)** | | |
| --- | --- | --- | --- | --- |
| **Total (n=1045)** | **Length of repeats (No of individuals)** | **AAAAG** | **AAAGG** | **AAGGG** |
| **Single Allele amplification (n=561)** | 5-15, 21-63 repeats | - | - | - |
| **Heterozygous (n=248)** |  |  |  |  |
| **No amplification (? Homozygous expansion); n=236^#^** | - | 1(A4G), 2 (A4G/A3G2), 2(A4G/A2G3) | 1(A3G2), 1(A3G2/A2G3), 2(A3G2/A4G) | 0(A2G2), 2(A4G/A3G2/A2G3) |

c. CMT

|  | **Flanking-PCR** | **Number of individuals with repeat expansions (Repeat-Primed PCR)** | | |
| --- | --- | --- | --- | --- |
| **Total (n= 93)** | **Length of repeats (No of individuals)** | **AAAAG** | **AAAGG** | **AAGGG** |
| **Single Allele amplification (n=38)** | 9-14, cases with repeats 41, 45 and 56 | - | - | - |
| **Heterozygous (n=17)** |  |  |  |  |
| **No amplification (? Homozygous expansion); n=38^#^** | - | NA(A4G), NA (A4G/A3G2), 0(A4G/A2G3) | NA(A3G2), 3(A3G2/A2G3) | 2(A2G3), 4(A4G/A3G2/A2G3) |

d. SCA12

|  | **Flanking-PCR** | **Number of individuals with repeat expansions (Repeat-Primed PCR)** | | |
| --- | --- | --- | --- | --- |
| **Total (n=87)** | **Length of repeats (No of individuals)** | **AAAAG** | **AAAGG** | **AAGGG** |
| **Single Allele amplification (n=31)** | 4-15, 33-111 repeats | - | - | - |
| **Heterozygous (n=49)** |  |  |  |  |
| **No amplification (? Homozygous expansion); n=7^#^** | - | 4 (A4G), 4 (A4G/A3G2), 0(A4G/A2G3) | 0(A3G2), 0(A3G2/A2G3) | 1(A2G3), 0(A4G/A3G2/A2G3) |

Table 2: Comparison between control and different disease groups with respect to allele distribution according to repeat number. (Number of cases corresponding to each studied group represents those whose RFC1-TNR size was estimated using flanking PCR and are normal)

|  | **P value (Chi square)** | | | |
| --- | --- | --- | --- | --- |
| **Repeat Size Category** | **Control (460)** | **Ataxia (809)** | **SCA12 (80)** | **CMT**  **(57)** |
| **SN** | **0.0001 (212.762)** | **0.0001 (762.841)** | **0.0001 (36.31)** | **0.0001 (69.17)** |
| **LN** | 0.1 (3.71) | **0.03 (4.9)** | 0.1 (2.2) | 0.6 (0.23) |
| **IA** | **0.0001 (240.772)** | **0.0001 (760.267)** | **0.0001 (31.39)** | **0.0001 (67.43)** |

Table 3: Comparison of each repeat size category at RFC1 locus in IndiGen control vs disease (including all cases) group using genotype count. (Number of cases corresponding to each studied group represents those whose RFC1-TNR size was estimated using flanking PCR and are normal)

|  | **P value (Chi Square Value)** | | | |
| --- | --- | --- | --- | --- |
| **Genotype Count** | **Controls (460)** | **Ataxia (809)** | **SCA12 (80)** | **CMT (57)** |
| **SN\|SN** | **<0.0001 (203.59)** | **0.0001 (654.9)** | **0.0001 (52.1)** | **0.0001 (61.89)** |
| **SN\|LN** | 0.57 (0.33) | **0.018 (5.6)** | 0.67 (0.18) | 0.41 (0.68) |
| **SN\|IA** | **<0.0001 (90.6)** | **<0.0001 (313.6)** | **0.0001 (28.4)** | **0.0001 (23.85)** |
| **LN\|IA** | 0.34 (0.9) | **0.019 (5.5)** | 0.8 (0.04) | 0.47 (0.52) |
| **IA\|IA** | **0.0001 (57.03)** | **<0.0001 (180.77)** | 0.13 (2.25) | **0.0001 (14.97)** |
| **LN\|LN** | **0.03 (4.5)** | 0.66 (0.2) | 0.46 (0.55) | 0.36 (0.8) |

Table 4: P value for association between repeat number and repeat type in IndiGen population. (AAAAG repeat as reference/control)

|  | **P value (Chi Square)** | | | | | |
| --- | --- | --- | --- | --- | --- | --- |
| **Repeat Motifs** | **IA\|IA** | **LN\|IA** | **LN\|LN** | **SN\|IA** | **SN\|LN** | **SN\|SN** |
| **AAAAGG** | 0.3492 (0.9) | - | - | 0.1108 (2.5) | - | - |
| **AAAGG** | **0.0308 (4.7)** | 0.3638 (0.8) | - | 0.2621 (1.3) | - | **0.0114 (6.4)** |
| **AAAGGG** | 0.728 (0.1) | - | - | 0.6719 (0.2) | **<0.0001 (16.2)** | 0.6844 (0.2) |
| **AACGG** | 0.509 (0.4) | - | **<0.0001 (57.8)** | 0.3291 (1) | - | **0.0007 (11.6)** |
| **AAGAG** | 0.0909 (2.9) | - | - | 0.0586 (3.6) | **0.0076 (7.1)** | - |
| **AAGGC** | 0.7695 (0.1) | - | **<0.00011 (87.2)** | 0.3011 (1.1) | **<0.00011 (57.7)** | - |
| **AAGGG** | 0.9155 (0) | - | - | 0.7292 (0.1) | - | - |
| **AGAGG** | 0.7695 (0.1) | - | - | 0.7512 (0.1) | **<0.0001 (57.7)** | - |
| **AAAAG/AAAGG** | 0.1966 (1.7) | 0.0603 (3.5) | - | 0.1625 (2) | - | - |
| **AAAAG/AAGAG/AAAGGG** | 0.1745 (1.8) | - | - | 0.8248 (0) | - | - |
| **AAAAG/AAGGC** | 0.1745 (1.8) | - | - | 0.8248 (0) | - | - |
| **AAAAG/AAGGG** | 0.5334 (0.4) | - | - | 0.6549 (0.2) | - | - |
| **AAAAGG/AAAAG** | 0.1745 (1.8) | - | - | 0.8248 (0) | - | - |
| **AAAAGG/AAAAG/AAGAG** | - | **<0.00011 (87.2)** | - | 0.8248 (0) | - | - |
| **AAAGG/AAGGC** | 0.1745 (1.8) | - | - | 0.8248 (0) | - | - |
| **AAAGGG/AAAAG** | **<0.0001 (34.2)** | 0.2646 (1.2) | - | **<0.0001 (31.8)** | - | 0.7791 (0.1) |
| **AAAGGG/AAAAG/AAAGG** | 0.1745 (1.8) | - | - | 0.8248 (0) | - | - |
| **AAAGGG/AAAGG** | 0.6586 (0.2) | **<0.0001 (42.9)** | - | 0.3011 (1.1) | - | - |
| **AAAGGG/AAGGG** | 0.0968 (2.8) | - | - | 0.1433 (2.1) | - | - |
| **AAAGGG/AGAGG** | - | - | - | 0.8248 (0) | **<0.0001 (116.7)** | - |
| **AACGG/AAAAG** | 0.1745 (1.8) | - | - | 0.8248 (0) | - | - |
| **AACGG/AAAGGG** | 0.1745 (1.8) | - | - | 0.8248 (0) | - | - |
| **AAGAC/AAAGGG** | 0.1745 (1.8) | - | - | 0.8248 (0) | - | - |
| **AAGAG/AAAAG** | 0.0021 (9.5) | - | - | 0.0063 (7.5) | - | - |
| **AAGAG/AAAGG** | 0.0968 (2.8) | - | - | 0.1433 (2.1) | - | - |
| **AAGAG/AAAGGG** | 0.6586 (0.2) | - | - | 0.7512 (0.1) | - | - |

Table 5: Primer sequence used for SNaPshot and sequencing for risk haplotype assessment in cases.

| **Primer ID** | **Seq (5'-> 3')** |
| --- | --- |
| rs2066790_IP | AATTCATGCAAAATTTAAAATAC |
| rs11096992_IP | CTCCTGTTAATATTAGTGAATGG |
| rs17584703_IP | ATTAAATCTCAGTGAAATTCAAA |
| rs6844176_IP | CTCCAAGTCTCAAGCTTCCAAAA |
| rs2066790_FP | TTCCCATAATCTGCCCTGAG |
| rs2066790_RP | CCTTGGGACCTTCTCGATTT |
| rs11096992_FP | CCAAGAATAAGCCTTTATCACCA |
| rs11096992_RP | ATGATCTTTTCCCGCTTTCA |
| rs17584703_FP | GCCTCCAGAGATGGGAAATA |
| rs17584703_RP | GACCTCACGGCTCTTCAAAC |
| rs6844176_FP | ACCCACATCGATGCAGTTTT |
| rs6844176_RP | GTCTTGAAGATGAAGAACGTGG |
